# Predictors of health related quality of life among diabetic patients on follow up at Nekemte Specialized Hospital, Western Ethiopia: A cross sectional study

**DOI:** 10.1101/19013516

**Authors:** Bikila Regassa Feyisa, Mekdes Tigistu Yilma, Belachew Etana Tolessa

## Abstract

**Objective:** To assess health related quality of life (HRQoL) and its predictors among diabetic patients on follow up at Nekemte Specialized Hospital (NSH) in Western Ethiopia.

**Design, setting and participants:** This facility based cross sectional study was conducted among 224 diabetic patients on follow up at one of the public hospitals in western Ethiopia.

**Main outcome measured:** HRQoL was measured by using the Medical Outcome Study 36-item Short Form Health Survey from 15^th^ April to 5^th^ June. Structured questionnaire was used for data the collection from participants selected by systematic random sampling. Multiple linear regression was used for final model.

**Result:** A total of 215 diabetic patients were involved in the study with the response rate of 96%. The mean score of the overall HRQoL of the study participants was found to be 50.30 ± 18.08 with highest mean score in physical functioning and lowest mean score in general health domain. Age, education status, history of smoking, feeling of stigmatized and Body mass index (BMI) were inversely associated while being male, being married, absence of co morbidity and absence of chronic complications related to diabetes mellitus were found to be positively associated with overall HRQoL.

**Conclusion:** The overall HRQoL of diabetic patients on follow up at the study area was found to be moderate. General health, mental health, bodily pain and vitality were the most affected domains. Both the mental and physical components need to be considered when caring the diabetic patients on follow up beyond provision of treatment.

**Strength and limitations of this study:**

- The study was the first of its kind in assessing the predictors of health-related quality of life (HRQoL) among both diabetes mellitus type I and type II in Ethiopia.
- The tools used was validated across different cultures
- Eight domains of HRQoL and two component summary scores were used to make the measurement more specific.
- The cross sectional nature of the study design made the result of the study difficult to identify either the cause or the effect comes first.

## Introduction

Diabetes Mellitus (DM) is defined as a metabolic disorder of numerous etiology which is characterized by chronic high blood glucose level (>126 mg/dl for fasting blood sugar and /or a 2-hour postprandial glucose of ≥200 mg/dl or if the individual had symptoms of diabetes and a random plasma glucose ≥200 mg/dl (confirmed by repeat testing) with disturbances of carbohydrate, fat and protein metabolism resulting from problems encountered in either insulin secretion problems (in case of type 1 diabetes mellitus or insulin action problem (type 2 diabetes mellitus) or both [1-3].

Diabetes mellitus is becoming a confronting problem of the time that have a considerable impact on health status and quality of life. It is considered an urgent public health problem because it has a pandemic potential, which can influence the HRQoL negatively [2, 15, 16].

Worldwide, 8.8 % of adults aged 20-79 years had diabetes in 2015 which was projected to reach 366 million in 2030 and 642 million (one in ten adults) by 2040, among which about 75% lived in low- and middle-income countries. Globally about 5 million deaths were attributable to diabetes in the 20–99 years’ age range [4, 6].

There is now a consensus that the health of general population cannot be well characterized from the analyses of mortality and morbidity statistics alone and that there is also a need to consider health in terms of people’s assessment of their sense of wellbeing and ability to perform social roles [7, 8].

Health-related quality of life (HRQoL) is a multidimensional concept that focuses on the impact of illness and treatment on patients, and it can measure patients’ perceptions of illness and treatment, their perceived needs for healthcare providers and their preferences for treatment and outcomes of the disease [9,10].

Health related quality of life is not only concerns subjective but also objective measurements of the individual with certain conditions. It has been defined as “an overall general well-being that comprises objective descriptors and subjective evaluations of physical, material, social, and emotional well-being together with the extent of personal development and purposeful activity, all weighted by a personal set of values [11].

Diabetes Mellitus negatively influences HRQoL. This negative influence affects many aspects of a person’s life, including the psychological impact of being chronically ill, dietary restrictions, changes in social life, symptoms of inadequate metabolic system, chronic complications and in due course lifelong infirmities [12, 13].

Studies have identified that diabetic patients’ HRQoL is decreased by different domains such as role limitation due to the disease, emotional disturbances, pain, and fatigability. Different factors related to health related quality of life among diabetic patients also affect both quality and quantity of life. These are socio demographics and economic status, behavioral, clinical and social related factors [18, 19].

Diabetes mellitus permanently changes the patient’s life style. Daily self-care, consisting of daily insulin injection or oral anti diabetic agents, self-monitoring of blood glucose and diabetic recommended diet has an impact on HRQoL. Moreover, the acute and chronic complications which might develop in due course affect the patients HRQoL [20]. In Ethiopia, even though programs have been launched regarding the chronic diseases management, prevention, screening, diagnosis, treatment and care, little is emphasized on factors that affect the HRQoL among the patients [21]. The guideline lacks the specific areas of HRQoL dimension which is affected by the disease. The existing articles and reviews have tried to indicate the epidemiology, complications, therapies, comparisons of treatments and health strategies but the data regarding associated factors of HRQoL and how much it is actually affected by the condition is scarce [20].

Therefore, this study was designed to predict the level of health related quality of life and factors associated with it among diabetic patients in Nekemte Specialized Hospital.

## Research Design and Method

### Study Design and setting

Facility based cross sectional study design was employed from 15^th^ April to 5^th^ June, 2019. The study was conducted among diabetic patients on follow up at Nekemte Specialized Hospital (NSH) which is found in Nekemte City and located to Western Ethiopia which is 331 km away from the capital city of Ethiopia, Addis Ababa.

Diabetic follow up and care services started separately as chronic diseases clinic at the Hospital in 2010 with 96 cases. According to the unpublished report of NSH taken on the 1^st^February, 2019,five hundred ninety one diabetic patients of both type 1 and type 2 were on follow up at the chronic disease clinic.

### Eligibility criteria

All known type I and type II DM patients who have been on follow up for at least one year duration and age greater than 18 years at NSH were included while diabetic patients who were seriously ill and could not respond to the interview were excluded from the study.

### Sample size and sampling procedure

The sample size was determined assuming a normally distributed independent mean, taking mean age value with standard deviation of 15.208 from previous study, 95% CI (Zα/2=1.96) and 5% marginal error. After calculating correction formula and adding 5% non response rate, the final sample size became 224.

Systematic random sampling was used to select the study participants. The sampling interval was developed from the diabetic identification number of the patients from the registry notebook and calculated by dividing the total number of diabetic patients on follow up by the calculated sample size.

### Data collection procedure

Data was collected using interviewer administered structured questionnaire which was adopted from the WHOQOL-BREF tool. The English version of the questionnaire was translated to Afaan Oromoo (local language) and translated back to English by other language experts to check its consistency. Four data collectors and one supervisor were recruited.

### Instruments

The instrument consists of the WHO SF-36 item questionnaires adopted from WHOQOL –BREF instrument and socio demographic and economic profiles. The SF-36 items consists of 36 questions containing physical functioning (10 items), role limitation due to physical health (4 items), body pain (2 items),vitality (4 items),social functioning (2 items), role limitation due to emotional problem (3 items), mental health (5 items) and general health (5 items).

### Data processing analysis

Each item of SF-36 was scored on linear scale and the negatively worded questions were inversely coded before any attempt of analysis. The score of each domain was obtained by summation of the corresponding items. The scores were then be linearly transformed on 0-100 scale. Mean scores were then be adjusted to make the domain scores comparable with the scores used in the WHOQoL-100 (Lower scores denote lower quality of life).

Summary scores on two subscales, the Physical Component Score (PCS) and Mental Component Score (MCS) derived from principal component factor analysis (PCA).

Dummy variables were created for categorical variables that have more than two categories like marital status, educational status, age category and the drug regimen.

For the internal consistency reliability of the SF-36 items, Cronbach’s alpha was checked and found to be 0.876 which was in the acceptable level.

Beta (β-Coefficient) was used to interpret the strength of predictors of HRQoL. The degree of association between pairs of variables was measured by Pearson’s correlation coefficient (r).The independent variables at P < 0.05 were considered as statistically significant. Multicollinearity was checked using variance inflation factors (VIFs). All covariates had value of VIF < 10 which is tolerable.

## Results

### Socio demographic characteristics

A total of 215 diabetic patients on follow up at NSH included in the analysis with response rate of 96%. Among the total respondents, 122 (56.7%) of them were males with mean age of 41.60 years with standard deviation (SD) of ±15.42and majority, 141 (65.6%) of them were from urban resident. Regarding the marital status, more than half, 146 (67.9%) of the total respondents were married among which males accounted for 58.9%% and majority, 198 (92.10%) of them were Oromo. The mean family size of the respondents was 4.83 with SD of±1.55. Thirty eight (17.70%) of the study participants could not read and write and only 52 (24.20%) of them were employed either at government and/or nongovernmental organizations **(Table 1)**.

**Table 1:** Socio demographic and socio economic characteristics of diabetic patients on follow up at NSH, Nekemte, East Wollega, West Ethiopia, 15th April -5th June, 2019, (n=215).

| Variables | Frequency (n=215) | Percentage |
| --- | --- | --- |
| **Sex** |  |  |
| Male | 122 | 56.7 |
| Female | 93 | 43.3 |
| **Mean age in year** | 41.60 (± SD 15.42) | |
| **Residence** |  |  |
| Urban | 141 | 65.6 |
| Rural | 74 | 34.4 |
| **Marital status** |  |  |
| Married | 146 | 67.9 |
| Single | 42 | 19.5 |
| Divorced | 6 | 2.8 |
| Widowed | 21 | 9.8 |
| **Ethnicity** |  |  |
| Oromo | 198 | 92.1 |
| Amhara | 9 | 4.2 |
| Guraghe | 5 | 2.3 |
| Other<sup>a</sup> | 3 | 1.4 |
| **Educational status** |  |  |
| Cannot read and write | 38 | 17.7 |
| Grade 1-8 | 63 | 29.3 |
| Grade 9-12 | 50 | 23.3 |
| College/University | 64 | 29.8 |
| **Occupation** |  |  |
| Government/NGO employee | 52 | 24.2 |
| Merchant | 23 | 10.7 |
| Farmer | 41 | 19.1 |
| Housewife | 46 | 21.4 |
| Retired | 23 | 10.7 |
| Other<sup>b</sup> | 30 | 14 |
| **Economic status (Wealth index)** |  |  |
| Poorest | 43 | 20 |
| Poor | 38 | 17.7 |
| Medium | 46 | 21.4 |
| Wealthy | 45 | 20.9 |
| Wealthiest | 43 | 20 |

<sup>a</sup>Other (Tigre, Silte) <sup>b</sup>Other (Student, carpenter)

### Medical history and health condition

More than half, 125 (58.1%) of the study participants had type 2 diabetes mellitus. Almost half, 108 (50.2%) of them had been diagnosed within last five years. Regarding treatment they were taking, 99 (46%) of the patients were using only insulin followed by 93 (43.3%) and 23 (10.7%) of them were using oral hypoglycemic agents and both insulin and oral hypoglycemic agents respectively.

Regarding the co morbidity status, nearly half, 103 (47.9%) of the study participants were co morbid with hypertension which accounts to 85 (82.5%) cases. Sixty three (29.3%) of the study participants had diabetes related acute complication in which 40 (63.5%), 20 (31.7%) and 3 (4.8%) constituted DKA, Hypoglycemia and non ketotic hyperosmolar state respectively. Sixty nine of the study subjects had diabetes related chronic complications which accounted around 32.1% where diabetic neuropathy (including foot ulcer, peripheral pain, and gangrene) covered almost half, 49.3% as illustrated in **table 2**.

**Table 2:** Medical history and health condition of diabetic patients on follow up at NSH, Nekemte, East Wollega, West Ethiopia, 15th April -5th June, 2019, (n=215).

| Variable | Frequency (n) | Percent(%) |
| --- | --- | --- |
| **Duration of DM (years)** |  |  |
| <5 | 108 | 50.20 |
| 6-10 | 59 | 27.40 |
| 11-15 | 32 | 14.90 |
| >15 | 16 | 7.40 |
| **Drug regimen** |  |  |
| Insulin only | 99 | 46 |
| Oral hypoglycemic agents | 93 | 43.3 |
| Insulin and oral hypoglycemic agents | 23 | 10.7 |
| **Presence of co morbidity** |  |  |
| Yes | 103 | 47.9 |
| No | 112 | 52.1 |
| **Presence of diabetic related chronic complication** |  |  |
| Yes | 69 | 32.1 |
| No | 146 | 67.9 |
| **Type of chronic complication** |  |  |
| Diabetic neuropathy | 34 | 49.30 |
| Diabetic Retinopathy | 19 | 27.50 |
| Diabetic nephropathy | 14 | 20.30 |
| Other | 2 | 2.80 |
| **Body mass index (BMI) (kg/m<sup>2</sup>)** |  |  |
| <18.5 | 5 | 2.30 |
| 18.5-25 | 122 | 56.70 |
| 25-30 | 80 | 37.20 |
| >30 | 8 | 3.7 |

### Health related quality of life of the study participants

Among the eight domains of HRQoL, the study participants scored highest (63.19±34.36) and lowest (30.21 ±22.95) mean score on physical functioning and general health domain respectively. When analyzing the HRQoL by domains general health, mental health, bodily pain and vitality had mean score below 50 indicating that they were the most affected domains among the diabetic patients. The transformed mean score of the overall HRQoL of the study participants was found to be 50.30 ± 18.08 with the minimum and maximum score of 16.38 and 79.13 respectively (**Table 3**).

**Table 3:** The eight domains of HRQoL, the overall HRQoL and the two component scores of HRQoL with their mean score of diabetic patients at NSH, East Wollega, West Ethiopia, 15th April -5th June, 2019,(n=215)

| **Domains of HRQoL, Overall HRQoL, PCS and MCS** | **Mean** | **SD** | **Minimum score (%)** | **Maximum score (%)** |
| --- | --- | --- | --- | --- |
| Physical Functioning (PF) | 63.19 | 34.36 | 0 | 100 |
| Role limitation due to physical health (RP) | 53.37 | 44.8 | 0 | 100 |
| Role limitation due to emotional problem (RE) | 52.71 | 45.82 | 0 | 100 |
| Energy/Fatigue (VT) | 48.47 | 7.78 | 20 | 75 |
| Emotional wellbeing (MH) | 49.84 | 8.02 | 20 | 72 |
| Social Functioning (SF) | 56.04 | 30.13 | 0 | 100 |
| Bodily Pain (BP) | 48.60 | 11 | 12.5 | 80 |
| General Health (GH) | 30.21 | 22.95 | 0 | 95 |
| Overall HRQoL | 50.30 | 18.08 | 16.38 | 79.13 |
| Physical Component Score (PCS) | 48.84 | 21.87 | 10 | 87.50 |
| Mental Component Score (MCS) | 51.77 | 16.72 | 19.75 | 80.75 |

**Table 4:** Multiple linear regression analysis of diabetic patients on follow up at NSH, East Wollega, West Ethiopia, 15th April -5th June, 2019,(n=215)

| Variables | Unstandardized Coefficient |  | Standardized Coefficient | 95% CI |  | P-value |
| --- | --- | --- | --- | --- | --- | --- |
| | B | SE | $\beta$ | Lower | Upper | |
| **(Constant)** | 69.41 | 10.31 |  | 49.16 | 89.74 | 0.000 |
| **Sex** |  |  |  |  |  |  |
| Female | 1 | 1 | 1 | 1 | 1 | 1 |
| Male | 5.23 | 2.11 | 0.14 | 1.10 | 9.36 | **0.013\*** |
| **Age** | -0.25 | 0.08 | -0.20 | -0.43 | -0.07 | **0.007\*** |
| **Marital status** |  |  |  |  |  |  |
| Single | 1 | 1 | 1 | 1 | 1 | 1 |
| Married | 5.30 | 2.69 | 0.11 | 0.88 | 10.52 | **0.046\*** |
| Divorced | -4.60 | 5.26 | -0.04 | -14.98 | 5.78 | 0.38 |
| Widowed | -4.07 | 3.42 | -0.07 | -10.81 | 2.67 | 0.24 |
| **Educational status** |  |  |  |  |  |  |
| Cannot read and write | -8.81 | 3.06 | -0.19 | -14.88 | -2.82 | **0.004\*** |
| Grade 1-8 | -2.94 | 2.49 | -0.07 | -7.84 | 1.97 | 0.24 |
| Grade 9-12 | 0.04 | 2.62 | 0.001 | -5.13 | 5.22 | 0.98 |
| College and above | 1 | 1 | 1 | 1 | 1 | 1 |
| **Smoking history** |  |  |  |  |  |  |
| Yes | -9.03 | 2.66 | -0.21 | -15.23 | -4.69 | **0.001\*\*** |
| No | 1 | 1 | 1 | 1 | 1 | 1 |
| **Feeling of stigmatized** |  |  |  |  |  |  |
| Yes | -5.25 | 1.89 | -0.15 | -8.94 | -1.56 | **0.005\*** |
| No | 1 | 1 | 1 | 1 | 1 | 1 |
| **Co morbidity status** |  |  |  |  |  |  |
| Yes | 1 | 1 | 1 | 1 | 1 | 1 |
| No | 6.05 | 2.18 | 0.16 | 1.78 | 10.33 | **0.006\*** |
| **Chronic complication status** |  |  |  |  |  |  |
| Yes | 1 | 1 | 1 | 1 | 1 | 1 |
| No | 6.04 | 2.28 | 0.11 | 1.54 | 10.53 | **0.009\*** |
| **BMI** | -3.56 | 1.71 | -0.12 | -6.94 | -0.18 | **0.040\*** |
| **DM Duration** | 0.15 | 1.10 | 0.01 | -2.02 | 2.33 | 0.89 |
| **Types of DM** |  |  |  |  |  |  |
| Type 1 | 1 | 1 | 1 | 1 | 1 | 1 |
| Type 2 | 4.45 | 2.39 | 0.12 | -0.24 | 9.17 | 0.064 |
| **Drug regimen** |  |  |  |  |  |  |
| Insulin only | 1 | 1 | 1 | 1 | 1 | 1 |
| OHA | -5.66 | 3.46 | -0.16 | -12.48 | 1.15 | 0.103 |
| Both | -1.42 | 4.02 | -0.02 | -9.35 | 6.51 | 0.72 |

Dependent Variable: Overall health related quality of life.

Two component scores of the HRQoL was also generated by PCA with the total variance explained 66.77%. The higher mean score was found for the mental component score (51.77±16.72) with the maximum score of 80.75.

### Predictors of health related quality of life of diabetic patients

The multiple linear regression model indicated that a unit increase in age would likely decrease health related quality of life of diabetic patients by 0.25 (*β*=-0.25, 95% CI, -0.43.55, -0.07, p=0.007) controlling all other independent variables.

Males had about five times better HRQoL when compared to their counter parts (*β*=5.23, 95% CI, 1.10-9.36, p=0.013). As for marital status, those who married had about five times better HRQoL when compared to those who were single controlling for all other independent variables (*β*=5.30, 95% CI,0.88-10.52. P=0.046).

Regarding the educational level the respondents achieved, those who were unable to read and write were about 9 times lower HRQoL (*β*=-8.81, 95% CI,-14.88 to -2.82, P=0.004) when compared to those who achieved college and above after controlling all other predictors.

History of smoking was found to affect the HRQoL status of the diabetic patients. Diabetic patients who had history of smoking had nine units times lower HRQoL (*β*=-9.03, 95% CI, -15.23--4.69, P<0.001) when compared to their counter parts. In the same way, feeling of stigmatized because of being diabetic patient would likely decrease HRQoL by 5.25 units (*β*=-5.25, 95% CI, -8.94 to -1.56, P=0.005) compared to their counterparts.

Absence of co morbid conditions and chronic complications related with diabetes mellitus was found to increase HRQoL when compared to their counter parts. In both cases, those who had not the condition had about six units better HRQoL than their counterparts. As for BMI, the increase in one unit of BMI would likely decrease the HRQoL by 3.56 units (*β*=-3.56, 95% CI,-6.94--0.18, P=0.009) **Table 4**

## Discussion

This study aimed to assess the overall health related quality of life with its domains and the predictors among diabetic patients of type I and type II in NSH.

According to this finding, the transformed overall mean score of the HRQoL was found to be moderate. The highest domain mean score was physical functioning. Domains of general health, mental health, bodily pain and vitality had mean score below the average. This result was lower when compared to the study result conducted in Saudi Arabia, Malaysia and Emirati people [17, 30, 34]. This discrepancy might be due to the difference in the socioeconomic status of the patients and the cultural difference across different regions. But it was congruent with the study result from Felege Hiwot Referral Hospital; Ethiopia [26].This might be because of the possible similarity of the socioeconomic status of the study areas where the study participants shared almost the same life style and cultural perspectives.

The physical and mental component mean score from the current study was comparable with the study from Denmark but higher than the study result reported from Greece [22, 24]. However, another study from Tahran Hospital indicated that the physical and mental component mean score was relatively higher than that of the present study. The possible explanation for this difference could be the cultural and socio demographic difference of the patients across different study areas. The subjective nature of the HRQoL and the component measurement across different patients might also indicate the differences HRQoL concerns subjective evaluations and also objective descriptors [10, 11].

In this study sex, age, marital status, educational level were among the socio demographic factors that had significant association with the health related quality of life with the diabetic patients. For instance, unable to read and write was inversely associated with the HRQoL indicating that diabetic patients who cannot read and write have lower understanding about the disease, the complication and the treatment as well as unable to make decision on better self care. The study disagreed with the study result from Greece where sex and educational level had not significant association with health related quality of life [10]. This difference could be because of the difference in socio demographic status of the patients.

As age increases the HRQoL of the diabetic patients would be decreased. This report was also noted in similar studies conducted in Greece, and Tehran Hospital, Iran [11, 25]. This might be because of the physiological alteration of the patients as they got older. Older individuals are mostly limited in physical activities, coping up the pain intensity and relief from pain [25]. However, findings from other parts of Ethiopia, South Africa and the Nordic countries were inconsistence with the current study result where age has no association with HRQoL [26,27, 29].

Married diabetic patients had higher HRQoL when compared to the single patients. The finding from the other part of Ethiopia also agreed with the current study result [26]. The possible explanation for this could be because of the married patients might be psychologically stable and have better social interaction in relation to those who are single.

The study identified that there was a gender difference in the mean score of the overall HRQoL. Male diabetic patients had higher HRQoL mean score when compared to the female patients with the highest domain score of physical functioning and lower in general health in both sexes. This goes in line with other literatures indicated that women got worse HRQoL than males [11, 28]. But it contradicted with other studies conducted in India, Tahran and Nordic countries [10, 25, 29]. This discrepancy could be due to the gender impact as most of the time women are treated inferiorly. They are less autonomous in giving decision on behalf of their rights.

Occupation and economic status were not found to be significantly associated with HRQoL of diabetic patients in this study. But, the finding from the same country, Ethiopia, identified as occupation was predictor for HRQoL of diabetic patients [26]. Another study also showed that economic status has significant association [12]. The discrepancy could be due to the methodological difference. Even though the patients share almost similar socioeconomic status, the previous studies analyzed the economic status just from mean annual income of the patients. However, since the patients’ way of disclosing their income level might not be accurate especially for rural residents, mean annual income might not be better way to forward conclusion. In this study, wealth index was computed.

In this study, patients who had history of smoking had decreased HRQoL when compared to their counter parts. This result was supported by the report from CDC and study from United States that indicated the direct impact of smoking altering the health condition of the diabetic patients and reduced their HRQoL [15, 31]. Smokers have more likely to have central fat accumulation than non-smokers, and smoking is known to induce insulin resistance and compensatory insulin secretion responses, which could explain the increased risk of diabetes in those who smoke [22]. But, it was not congruent with the study from South Africa where no association between cigarette smoking and HRQoL of diabetic patients were declared [27, 33]. This could likely be because of the effect of the sample size.

Both co morbidity condition and chronic complication related to diabetic was found to affect the HRQoL status which was similar with different studies conducted so far in Singapore, Arab Emirates, Saudi Arabia and Ethiopia [6, 17, 23, 26, 32]. This could be due to the fact that co morbid conditions which in this study like hypertension is another challenging that could put the patients in worrying conditions. Patients might seek health care for both or above diseases in which case they were emotionally diseased, the role due to emotional problem might be under question. All the domains of HRQoL directly or indirectly would be affected. In another way, those who developed chronic complications were also live under the double crisis. In one way, they felt unhappy of being diabetic patient and in other way they would be under the psychological, physical, emotional, social and spiritual agony.

However, in Greece presence of complications of diabetes mellitus had no significant association with HRQoL of patients which is different from the current study finding [11].This might be due to the difference in better access to care in the management of diabetes patients.

In this study, the increase in BMI affected HRQoL negatively. But in South Africa and Greece there was no significant association between BMI and HRQoL [11, 27]. This discrepancy could be due to the difference in the diabetic patients’ knowledge gap and practice regarding their life style modification.

Feeling of stigmatized of being diabetic patient inversely associated with HRQoL. This result goes in line with other literatures [18]. Because the diabetic patients are living under multiple restrictions, they would likely feel stigmatized in all aspects of their life. They are often restricted with regard to the amount, type and timing of food consumed. For example, eating mandatory foods at certain times, waiting for insulin to take effect before eating, etc. These restrictions may negatively affect an individual’s HRQoL and their interaction with people around them, in their social lives and in the work place.

The study had several limitations that have to be put in to consideration when used by other researchers. Since the study was a cross sectional study design, it is difficult to infer the cause effect relationship (temporal relation). The study was facility based which could not be generalized to all diabetic patients remained in the community. Face-to-face interview was conducted by considering the different level of education of the participants, which might lead to the social desirability bias and could overestimate the result. The effect of recall bias cannot be ruled out. The health care providers view towards the health related quality of life was not assessed.

In conclusion, the present study identified that the HRQoL of diabetic patients on follow up at NSH was moderate. Domains of general health, mental health, bodily pain and vitality were the most affected domains among the diabetic patients. Sex, age, education status, marital status, history of smoking status, BMI, feeling of stigma status, co morbidity status and diabetic related chronic complication status were predictors of health related quality of life identified in this study.

However, residence, economic status, occupation, type of diabetes, drug regimen and DM duration were not statistically significant predictors of health related quality of life.

There are several avenues for further researches based on the current finding. Longitudinal studies with larger sample size needs to be conducted in order to generalize the overall health related quality of life of diabetic patients at national level. Moreover, experimental and qualitative study design needs to be considered focusing the life style modification of the diabetic patients.

## Data Availability

Data could be found on demand from the email address of the corresponding author

## Compliance with ethical standards

### Funding

This work was financially supported by Wollega University with grant number WU/RD/256/2011.

### Ethical approval

The study received letter of approval from Research Ethics Review Committee (RERC) of Wollega University, DPH/0081/2011.

### Informed consent

Informed consent was obtained from all individual participants involved in the study.

### Authors’ contributions

All authors were contributed from the conception of the study to the final draft of the manuscript. Material preparation, data collection, analysis and interpretation of the result were carried out by Bikila Regassa, Mekdes Tigistu and Belachew Etana. The first draft of the manuscript was written by BR and reviewed by MT and BE. All authors read and approved the final manuscript.

## Acknowledgements

We are grateful to Wollega University, the study participants, data collectors and all other personnel who contribute their valuable input for the study to be accomplished.

## Annexes

Table showing correlation matrix of the eight domains among *diabetic patients on follow up at NSH, East Wollega, West Ethiopia, 15th April -5th June, 2019,(n=215)*

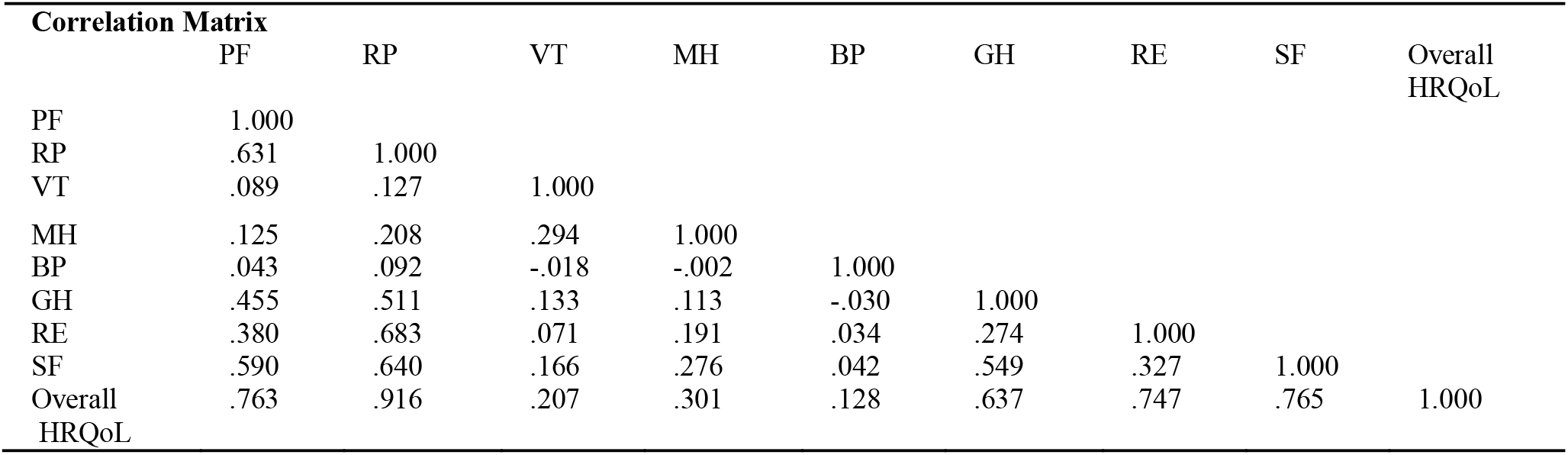

| **Correlation Matrix** |  |  |  |  |  |  |  |  |  |
| --- | --- | --- | --- | --- | --- | --- | --- | --- | --- |
|  | PF | RP | VT | MH | BP | GH | RE | SF | Overall HRQoL |
| PF | 1.000 |  |  |  |  |  |  |  |  |
| RP | .631 | 1.000 |  |  |  |  |  |  |  |
| VT | .089 | .127 | 1.000 |  |  |  |  |  |  |
| MH | .125 | .208 | .294 | 1.000 |  |  |  |  |  |
| BP | .043 | .092 | -.018 | -.002 | 1.000 |  |  |  |  |
| GH | .455 | .511 | .133 | .113 | -.030 | 1.000 |  |  |  |
| RE | .380 | .683 | .071 | .191 | .034 | .274 | 1.000 |  |  |
| SF | .590 | .640 | .166 | .276 | .042 | .549 | .327 | 1.000 |  |
| Overall HRQoL | .763 | .916 | .207 | .301 | .128 | .637 | .747 | .765 | 1.000 |

